# Evaluation of a food access program on inpatient nutrition, recovery, cost of care, and health related quality of life in selected public hospitals in Rwanda

**DOI:** 10.1101/2025.02.27.25323048

**Authors:** Stefanie L. Weiland, Edouard Musabanganji, Maïli Mia Gasakure, Linda Ntaganzwa

## Abstract

**Introduction:** Better nutrition is a priority in African countries like Rwanda with high prevalence of undernutrition and growing rates of obesity. Since public health insurance does not cover meals, vulnerable hospital inpatients are at risk of deficiency. We assessed the nutrition, health, and wellbeing effects of providing meals through the *Gemura* hospital feeding program.

**Methods:** We tested outcomes of the intervention in four public hospitals on inpatients (n=794). The primary outcome of nutritional status as body mass index (BMI) was measured at admission and discharge. Additional data collected included daily food intake, length of stay (LOS), patient costs of care, HRQOL using the EuroQol EQ-5D-3L tool, and patient satisfaction based on the Net Promoter Score (NPS).

**Results:** Mean weight in the *Gemura* group increased slightly (+0.6%), but the control group experienced a drop in average weight (-1.5%), with p=0.000. BMI distribution was improved generally in the *Gemura* group compared to controls, with the odds of being malnourished (BMI<18.5) at endline 9% greater in the control group (OR 1.09, p=0.000). LOS had weak association with study arm. Intervention group participants spent on average 40% less (10,985 RWF) on food and beverages than the control group (18,290 RWF) with high significance at p=0.000, while the control group spent less on total out of pocket costs of stay. Patient satisfaction and HRQOL scores were similar, except 40% less patients reported anxiety or depression in the *Gemura* group.

**Conclusion:** While further investigation is needed to assess recovery and patient-reported outcomes, improvements in nutrition, mental health, and out of pocket patient costs indicate the potential for the *Gemura* program to address nutrition gaps in hospital service delivery, to improve mental health, and to reduce economic barriers to care.

**What is already known on this topic:** Malnutrition has been associated with poor hospital outcomes, but few have studied access to nutritious food as a barrier, with scant evidence from intervention studies on inpatient food access in Sub-Saharan Africa, and none in Rwanda.

**What this study adds:** In this study, we found that providing three nutritious meals a day to hospital inpatients in Rwanda improved food intake, reduced the odds of hospital malnutrition, reduced patient out-of-pocket costs, and had a positive effect on patient perceptions of their mental health compared to a control group. This is the first intervention study of its kind in Rwanda.

**How this study might affect research, practice or policy:** In LMIC contexts of high malnutrition, these findings may help inform considerations of including hospital nutrition access programs in public health insurance schemes or other healthcare improvement initiatives.

## INTRODUCTION

The need for better nutrition, globally recognized through Sustainable Development Goal (SDG) 2, “End hunger, achieve food security and improved nutrition and promote sustainable agriculture,”^1^ is a high priority for Rwanda where 35.8% of the population is undernourished, the 6^th^ highest rate in the Sub-Saharan African (SSA) region.^2^ Nutrition also underpins several other SDGs and plays a major role in health and wellbeing (SDG3), including recovery from illness. Deficiencies enable or exacerbate a range of negative outcomes like immune system weaknesses and increased risk of infection, slower recovery and longer length of stay in hospital (LOS), increased treatment costs, higher mortality, higher readmission rates, missed socioeconomic opportunities, and decreased productivity.^3–13^ Lower socioeconomic status is in turn linked with lower nutritional status. Despite Rwanda’s nearly miraculous reduction in poverty rates and economic growth over the past two decades, malnutrition remains high, with rates of stunting remaining stagnant for at least a decade,^14^ and highest in rural and poor households which also experience high rates of food insecurity.^15^ At the same time, there are growing rates of overweight and obesity in Rwanda, indicating a double burden of malnutrition.

The period of time a patient is in the hospital is crucial for good nutrition, which has been shown to enable healing and recovery,^16–22^ but there is unfortunately scant data on hospital nutrition in SSA and specifically in Rwanda. Findings from the few published single-site observational studies on specific patient groups in a Kigali hospital show high prevalence of baseline malnutrition and associated negative outcomes, significant weight loss and increased malnutrition during hospitalization, as well as inadequate food intake.^23–26^

Patients from lower socioeconomic classes are likely at higher risk for hospital malnutrition— decreased nutritional status during hospitalization—since food provision is an extra cost not currently covered by government health insurance benefits. Many lower income patients experience challenges paying for food— 83% reporting a financial burden obtaining food during hospitalization in one study in Rwanda^26^—as meals from hospital canteens and external vendors can exceed insurance copays for treatment.^27^ The *Gemura* program of Solid’Africa aims to improve nutrition and recovery for these vulnerable patients through free provision of three nutritious meals a day during their stay in public hospitals in Rwanda.

The aim of this study was to test the effect of this feeding intervention on hospital inpatients’ health and wellbeing, assessing the primary outcome of nutrition during the hospital stay. Other objectives included examining the effect of the program on patient recovery during the hospital stay; costs of hospital care from a patient perspective; and patient satisfaction and health-related quality of life. As an intervention study, it provides unique outcome data on specific efforts to improve nutritional status, recovery, and patient experience through hospital feeding programs that can inform government and partner organization policies to improve health equity in Rwanda and beyond. Specifically, this study provides Solid’Africa and the Government of Rwanda essential data for decision-making on program continuity and scale-up.

## METHODS

We tested the effects of the *Gemura* program comparing differences in outcomes between the intervention group a control group in four selected public hospitals from urban and peri-urban areas, including a quasi-experimental difference-in-differences analysis. The intervention sites control sites were comparable in type of hospital in the healthcare system and in services to patients. The target sample (n=848), divided equally among sites, was statistically powered on the primary outcome of nutritional status (BMI) at a 95% confidence level, with further consideration for design effect (1:2), contingency (5%), and dropout (5%).

Participants were selected according to the existing procedures used by Solid’Africa and their partner hospitals to identify patients eligible for the program. This procedure involves hospital staff identifying patients’ socioeconomic vulnerability at intake, which is part of hospital social workers’ usual responsibilities. Among these vulnerable inpatients, the study included Rwandan children and adults over 5 years of age, admitted for any reason other than maternity delivery or specialized nutrition treatment, able to voluntarily feed themselves and stand on a scale for weight measurement. Eligible participants were enrolled upon informed consent as they were admitted from June – September 2024.

Data was collected at enrolment, during the hospital stay and at discharge, with no follow-up period. Food access exposure was measured through the *Gemura* program data at intervention sites and daily food intake surveys for all participants. Nutritional status was measured anthropometrically in terms of weight-for-height by body mass index (BMI = kg/m^2^) at baseline and endline, with BMI < 18.5 considered malnourished, 18.5-20.0 underweight, 20.1-24.9 normal weight, 25.0-29.9 overweight, and 30.0+ obese.^28^

Recovery during hospitalization was measured quantitatively as length of stay (LOS) in fractions of days as well as a qualitative survey of treatment outcome on discharge by the patient and their attending healthcare worker. Health-related quality of life was measured using the Kinyarwanda version of the EuroQol EQ-5D-3L health questionnaire consisting of five dimensions: mobility, self-care, usual activities, pain/discomfort, and anxiety/depression.^29–30^ Patient satisfaction was measured with a qualitative survey open question based on the Net Promoter Score (NPS).^31^ The survey question was developed with reference to the UK National Health Service’s (NHS) most recently updated patient survey guidance.^32^ Costs of care were measured from the patients’ perspective and estimated where appropriate in RWF and collected from both health facility records of patient treatment costs as well as from patients in their discharge survey to include money spent on food and transportation.

Additional descriptive data was collected from participants at baseline to be used to analyse drivers and confounders, including disease or condition, hospital ward/department, HIV status, baseline nutritional status, age, and sex. Sex was assigned by research assistants following external exam of body characteristics by healthcare workers during hospital intake.

### Intervention

The Solid’Africa *Gemura* intervention was administered as usual for study participants in the two intervention sites. Eligible hospital inpatients received three meals a day during their stay, as prescribed by hospital nutritionists for their specific conditions and ordered from Solid’Africa. The Solid’Africa team prepared the meals in their off-site kitchen and delivered them daily to the hospitals where the meals were distributed to the study participants. Meal caloric and nutritional content information is standardized except upon request. The normal meal is 575g of food consisting of 2300kCal with 300g of carbohydrates, 175g of protein, and 100g vitamins/minerals. In addition, Solid’Africa adjusts meal content per nutritionist request for specific needs like low salt/sugar, etc. The intervention was provided to more patients than were enrolled in the study, including pregnant women admitted for delivery, because of the organization’s vision for equity. For this reason, and because the intervention was purposively allocated to socioeconomically vulnerable inpatients, randomization was not employed.

### Analysis

Participant characteristics and outcome variables BMI, LOS, cost of treatment, patient satisfaction, and HRQOL descriptive statistics were calculated with 95% confidence intervals using IBM SPSS STATISTICS v.26 and STATA v.16.1 Relationships between outcomes and other variables were assessed using bivariate analysis with independent t-tests, and analysis of variance to evaluate the differences in group means at 5% of level of significance. The Pearson’s chi-square test was used to assess the independence between qualitative variables at a significance level of 5%. Fisher’s exact test was used as a complementary technique, specifically for categories with smaller (less than 5) expected frequencies. In addition to bivariate analysis, linear regression and kernel regression models were built for the quantitative outcome variables to include possible interactions between predictor variables and the intervention, and control for individual-level unobserved and time-invariant characteristics.

In addition, the intervention effect was analysed using a difference-in-differences (DID) analysis to estimate the change in outcomes between intervention and control groups: between groups with or without the intervention and before-and-after the intervention. We calculated DID = (B-A)-(D-C) for BMI and % weight change, comparing changes for participants receiving the *Gemura* intervention (B – A) with the change in outcomes for participants that did not receive the intervention (D – C), or the counterfactual.

The SQUIRE reporting guidelines for quality improvement in healthcare^33^ were used during this study.

### Ethical Considerations

Ethical approval for the study was obtained from the Rwanda National Ethics Committee (RNEC 80/2024) on 12 April 2024, the Ethics Committee of University Teaching Hospital of Kigali (CHUK) (EC/CHUK/122/2024) on 11 June 2024, and the Ethics Committee of University Teaching Hospital of Butare (CHUB) (REC/UTHB/133/2024) on 22 May 2024. Informed consent was obtained from all individuals whose data are included in the analysis. This study was also registered with Clinicaltrials.gov with identifier NCT06852885.

### Patient and Public Involvement

Patients were not directly involved in forming this research question, designing this study, or conducting this study.

## RESULTS

We aimed to enrol 848 participants across four hospitals and ultimately achieved n=918 participants, a response rate of 108.2%. Due to various factors such as deaths and transfers to other hospitals, the study concluded with n=794 participant datasets, representing 93.6% of the target sample, still sufficient for the reliability of the findings.

### Participant Characteristics

Overall, there were more males (58.2%) than females (41.8%), and an imbalance between study arms, with slightly more males than females in the control group (61.3% v. 54.4% male). The mean age of all participants was 40.8 years (SD=20.1, range: 5-95), with wide age diversity in the sample. Mean and median ages were quite similar across all sites, but the two larger university hospitals had slightly lower mean ages and children under 9 years in their sample. There was notable variation in geographic origin of participants across study sites, with all provinces represented (22% Kigali; 26% Northern; 18% Southern; 24% Western; 10% Eastern) among patients at CHUK, the university hospital located in Kigali, but predominantly local representation in other sites (90% Southern at CHUB; 72% Kigali at Masaka; 100% Eastern at Nyamata). The majority of participants (67.3%) were HIV negative, and HIV positivity was relatively low (2.6%) across all sites. However, a large proportion of patients at CHUB (81.9%) and to a lesser extent at Masaka (28.6%), reported “unknown” HIV status. At baseline, the mean weight was 55.4 kg (SD=14.3, range: 15-140), only marginally higher in the intervention group (55.9 kg) than in the control group (55.0 kg). Mean height was recorded at 161.9 cm (SD=14.7, range: 95-190), and was very similar among intervention (162.4 cm) and control (161.4 cm).

Patient diagnoses were grouped in categories, with distribution across study arms presented in Table 1. We found a significant relationship between diagnosis categories and study arms using Pearson’s chi-square (211.888, p < 0.001) and Fisher’s exact test (230.715, p < 0.001). Categories with higher frequencies include *Burns, puncture wounds, surgical procedures, and fractures* (31.6% in the control group vs. 13.6% in the intervention group) and *Infectious Diseases and Inflammatory Responses* (21.4% in the control group vs. 14.1% in the intervention group). This indicates that the control group had a significantly higher proportion of patients diagnosed with trauma-related and infectious conditions. Conversely, the *Not Classified* category accounted for 30.1% in the intervention group but only 3.4% in the control group, suggesting a distinct difference in the distribution of non-categorized conditions across the groups. The category *Non-Communicable Diseases* (NCDs) shows another notable difference, with the intervention group having a much higher proportion (13.6%) compared to the control group (4.4%). In contrast,

**Table 1.** Diagnosis categories across study arms.

| Diagnosis Categories | Control Group | Gemura Group | Total |
| --- | --- | --- | --- |
| Big Three (HIV, TB, and Malaria) | 12 (2.4%) | 21 (5.1%) | 33 (3.6%) |
| Non-Communicable Diseases (Diabetes, Hypertension, Coronary, Heart Disease) | 22 (4.4%) | 56 (13.6%) | 78 (8.6%) |
| Infectious Disease and Inflammatory Response (Pneumonia, Gastroenteritis, tuberculosis, measles, pertussis, etc.) | 107 (21.4%) | 58 (14.1%) | 165 (18.1%) |
| Respiratory Diseases | 10 (2.0%) | 9 (2.2%) | 19 (2.1%) |
| Women-Centred (Maternal health, vaginal bleeding, and fistula) | 51 (10.2%) | 2 (0.5%) | 53 (5.8%) |
| Burns, puncture wounds, surgical procedure and fractures | 158 (31.6%) | 56 (13.6%) | 214 (23.5%) |
| Nervous System, neurology, and mental health | 14 (2.8%) | 8 (1.9%) | 22 (2.4%) |
| Cancer, Tumour and Cyst/Mass | 56 (11.2%) | 35 (8.5%) | 91 (10.0%) |
| Haemorrhage (external and internal) | 2 (0.4%) | 0 (0.0%) | 2 (0.2%) |
| GI Tract | 20 (4.0%) | 17 (4.1%) | 37 (4.1%) |
| Immuno-related diseases, Blood related | 18 (3.6%) | 20 (4.9%) | 38 (4.2%) |
| Urinary System | 13 (2.6%) | 6 (1.5%) | 19 (2.1%) |
| Not Classified | 17 (3.4%) | 124 (30.1%) | 141 (15.5%) |
| **Total** | **500 (100.0%)** | **412 (100.0%)** | **912 (100.0%)** |

*Women-centred* conditions (i.e. gynaecological) appear predominantly in the control group (10.2%) compared to only 0.5% in the intervention group.

### Weight Change and BMI

The intervention group mean weight increased slightly (from 55.9 to 56.0 kg) during patients’ hospital stay, but the control group experienced a drop in mean weight (from 55.0 to 54.3 kg). Males and females in the intervention group experienced a positive mean weight change as a percent of baseline (+0.6% total), and both sexes in the control group experienced a drop in average weight (-1.5% total), with p=0.000 from the Mann-Whitney U test. Twenty percent (20.4%) of the *Gemura* intervention group lost more than 2% of body weight compared to 36.6% in controls.

Comparing BMI changes across study arms, presented in Table 2, reveals more nuanced differences. The baseline BMI results show a relatively similar distribution of BMI categories between the control and intervention groups. The endline BMI results reveal that the intervention group experienced notable effect, particularly among females, in improving BMI distribution compared to the control group. In the intervention group, a smaller proportion were classified as malnourished (25.4%) compared to controls (27.0%), and while the proportion of malnourished grew in the control group, it decreased in the *Gemura* group. Furthermore, the proportion of those in the normal BMI range increased in the *Gemura* group, but decreased in the control group, and the proportion of those in overweight and obese categories combined decreased in the *Gemura* group and increased in the control group.

**Table 2.** Baseline and endline BMI across study arms disaggregated by sex.

|  | Control Group |  |  |  |  |  | Gemura Group |  |  |  |  |  |
| --- | --- | --- | --- | --- | --- | --- | --- | --- | --- | --- | --- | --- |
|  | Baseline |  |  | Endline |  |  | Baseline |  |  | Endline |  |  |
|  | Male | Female | Total | Male | Female | Total | Male | Female | Total | Male | Female | Total |
|  | N (%) | N (%) | N (%) | N (%) | N (%) | N (%) | N (%) | N (%) | N (%) | N (%) | N (%) | N (%) |
| Malnourished:<br>BMI < 18.5 | 83 (27.9) | 37 (20.1) | 120 (24.9) | 93 (31.1) | 38 (20.4) | 131 (27.0) | 63 (28.8) | 46 (26.1) | 109 (27.6) | 47 (27.6) | 33 (22.8) | 80 (25.4) |
| Underweight:<br>BMI 18.5-19.9 | 52 (17.4) | 22 (12.0) | 74 (15.4) | 52 (17.4) | 28 (15.1) | 80 (16.5) | 34 (15.5) | 26 (14.8) | 60 (15.2) | 33 (19.4) | 24 (16.6) | 57 (18.1) |
| Normal: BMI<br>20-24.9 | 152 (51.0) | 95 (51.6) | 247 (51.2) | 144 (48.2) | 88 (47.3) | 232 (47.8) | 100 (45.7) | 71 (40.3) | 171 (43.3) | 76 (44.7) | 65 (44.8) | 141 (44.8) |
| Overweight:<br>BMI 25 – 29.9 | 10 (3.4) | 25 (13.6) | 35 (7.3) | 9 (3.0) | 30 (16.1) | 39 (8.0) | 19 (8.7) | 23 (13.1) | 42 (10.6) | 11 (6.5) | 15 (10.3) | 26 (8.3) |
| Obese:<br>BMI >30 | 1 (0.3) | 5 (2.7) | 6 (1.2) | 1 (0.3) | 2 (1.1) | 3 (0.6) | 3 (1.4) | 10 (5.7) | 13 (3.3) | 3 (1.8) | 8 (5.5) | 11 (3.5) |
| **Total** | **298 (100.0)** | **184 (100.0)** | **482 (100.0)** | **299 (100)** | **186 (100)** | **485 (100)** | **219 (100.0)** | **176 (100.0)** | **395 (100.0)** | **170 (100)** | **145 (100)** | **315 (100)** |

These results suggest that the intervention had a positive impact, particularly in reducing malnutrition. The proportion of malnourished increased in the control group relative to the *Gemura* group, with an odds ratio (OR) of 1.09, indicating 9% greater odds of being malnourished at endline in the control group. Using the Pearson’s chi-square test, we found the association between prevalence of malnutrition and study arm to be strong, at p=0.000.

During the analysis, we also found a significant association between diagnosis category and BMI with the Pearson’s chi-square test in both the *Gemura* intervention group (p<0.001) and in the control group (p=0.028) at baseline. At the endline however, this association persisted in the *Gemura* intervention group (p=0.017), but weakened in the control group (p=0.142). This data indicates that other factors besides diagnosis had a greater effect on BMI and that the *Gemura* program likely contributed to the sustained association between BMI and diagnosis.

### Difference-in-Differences (DID)

We used DID analysis to assess the effect of the feeding intervention on both BMI and weight change. While the analysis validated the differences among groups described above, the p-values from regression analysis of 0.496 and 0.573 shown in Table 3 indicate very little evidence that the *Gemura* program had a significant effect on BMI or weight.

**Table 3.**
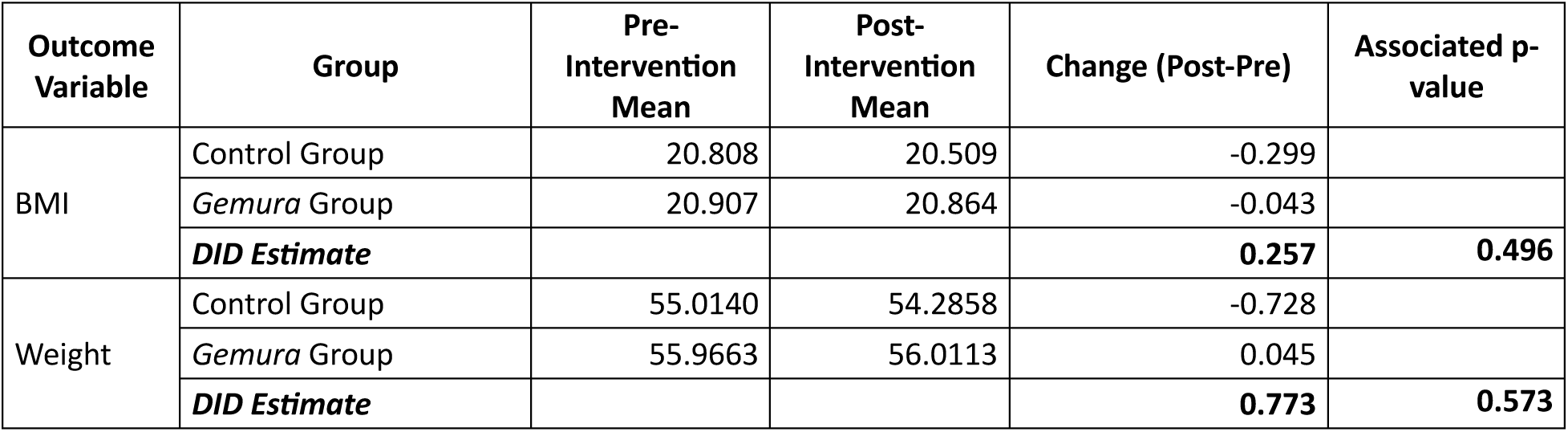
Difference-in-Differences effect estimates.

| Outcome Variable | Group | Pre-Intervention Mean | Post-Intervention Mean | Change (Post-Pre) | Associated p-value |
| --- | --- | --- | --- | --- | --- |
| BMI | Control Group | 20.808 | 20.509 | -0.299 |  |
|  | *Gemura* Group | 20.907 | 20.864 | -0.043 |  |
|  | **DID Estimate** |  |  | **0.257** | **0.496** |
| Weight | Control Group | 55.0140 | 54.2858 | -0.728 |  |
|  | *Gemura* Group | 55.9663 | 56.0113 | 0.045 |  |
|  | **DID Estimate** |  |  | **0.773** | **0.573** |

We then included additional patient characteristic variables in the regression model to compare with the estimated program effect (DID), shown in Table 4. The coefficients (B) and associated p-values for these variables provide insight into the factors influencing the outcomes of BMI and weight. For BMI, the magnitude of the program effect is much higher than the other variables, but the p-value of 0.312 is not statistically significant. Age and sex had the strongest evidence of effect, while province, LOS, and diagnosis also showed moderately strong evidence of effect. The other p-values imply that other, unmeasured factors may explain changes in BMI. Similarly for weight, the program effect was not statistically significant at p=0.475, while sex, province, age, and LOS had significant effects. The results suggest that other, unaccounted for factors may have influenced both BMI and weight change.

**Table 4.** Difference-in-Difference regression estimates with covariates for BMI and Weight.

|  | Weight |  |  |  | BMI |  |  |  |
| --- | --- | --- | --- | --- | --- | --- | --- | --- |
|  | Unstandardized Coefficients |  | Test statistic | p-value | Unstandardized Coefficients |  | Test statistic | p-value |
|  | B | Std. Err. |  |  | B | Std. Err. |  |  |
| (Constant) | 51.080 | 1.368 | 37.330 | 0.000 | 19.804 | 0.384 | 51.510 | 0.000 |
| Intervention | 0.130 | 1.067 | 0.122 | 0.903 | -0.275 | 0.299 | -0.918 | 0.359 |
| Post-Intervention | -0.789 | 0.847 | -0.932 | 0.351 | -0.304 | 0.237 | -1.280 | 0.201 |
| Diff-in-Diff (DID) | 0.962 | 1.347 | 0.715 | 0.475 | 0.382 | 0.378 | 1.011 | 0.312 |
| Sex | 2.458 | 0.676 | 3.638 | 0.000 | 1.264 | 0.190 | 6.670 | 0.000 |
| Province | -1.223 | 0.310 | -3.948 | 0.000 | -0.170 | 0.087 | -1.958 | 0.050 |
| Age | 0.175 | 0.017 | 10.549 | 0.000 | 0.018 | 0.005 | 3.929 | 0.000 |
| Length of stay (LOS) | -0.156 | 0.057 | -2.745 | 0.006 | -0.032 | 0.016 | -1.981 | 0.048 |
| Diagnosis | -0.051 | 0.093 | -0.545 | 0.586 | 0.060 | 0.026 | 2.289 | 0.022 |

### Secondary Outcomes

Food intake results from daily patient surveys showed a stark difference between control and intervention groups. In the control group, 70.3% reported eating "too little" during their hospital stay versus only 9.8% of patients in the *Gemura* group. The vast majority (87.8%) receiving the intervention indicated they had "enough" food, and a small percentage (2.4%) even reported eating "too much," a response absent in the control group. The difference between the groups was confirmed by the chi-square test result (p = 0.000), indicating that the feeding program was significantly associated with improved food intake among patients.

Recovery time results showed mean length of stay (LOS) was higher in the *Gemura* group (8.35 days) than in the control group (4.81 days), a significant difference with p=0.000. However, Kernel regression models did not show large mean effects of variables including age (10.254), diagnosis category (1.870), baseline BMI (1.807), province (0.626), sex (0.252), and study arms (0.250). Age showed modest effect, and diagnosis category and baseline BMI showed small effects, but were insufficient to account for the LOS difference between groups. Discharge surveys showed marginal differences in patient reported recovery outcome, with over 95% in both groups either agreeing or strongly agreeing with the statement: “*My treatment at the hospital was effective in making my condition better.”*

Results of patient out-of-pocket expense data analysis with the Mann-Whitney U test show that patients in the *Gemura* group spent considerably less on food and beverages (median= 6,000 RWF) while in hospital than the control group (median=15,000 RWF), with significance at p=0.000. The control group median total costs of hospital stay (29,360 RWF) were slightly lower than in the *Gemura* group (31,600 RWF), but with less significance at p=0.013, and the *Gemura* group mean total hospital cost per day (5,740 RWF) was lower than among controls (7,688 RWF).

For patient satisfaction, patients were asked a question based on the Net Promoter Score^34^ to indicate the likelihood of recommending similar care at this hospital to a friend or relative. Results showed the control group had a slightly higher mean score (8.85 out of 10) compared to the *Gemura* group (8.71), as well as more “promoters” (scores 9-10) and less “passives” (scores 7-8), indicating that overall satisfaction was higher in the control group. However, an independent t-test showed weak significance of the difference between groups with p=0.052.

For health-related quality of life, we used the EuroQoL EQ-5D-3L, in which patients assessed their health in five dimensions and three levels of response: no problems (1), some problems (2), extreme problems (3). Results showed scores were marginally better in mobility, selfcare, and usual activities dimensions in the control group. But higher percentages of the *Gemura* group reported no problems with pain/discomfort (53.6% versus 49.9%) or anxiety/depression (82.8% versus 71.6%) compared to controls. The second part of the questionnaire consisted of a visual analogue scale of 0 (the worst imaginable health) to 100 (the best imaginable health). Higher mean scores in the control group (84.2) indicate a better perceived quality of life than in the intervention group (74.1). The intervention group had greater variability in scores, with a standard deviation of 17.7 versus 10.7 in the control group, and a larger range of scores.

## DISCUSSION

We used patient data collected before, during, and at discharge from hospital treatment to assess the primary effects of the *Gemura* hospital feeding program on nutritional status, and secondary effects on recovery, patient costs of care, patient satisfaction, and health-related quality of life.

Our findings on food intake, weight change, and BMI show that there were statistically significant differences between the intervention and control groups, indicating the program had a positive effect on nutrition. Since the proportion of *Gemura* group patients reporting having eaten enough while in hospital was almost three times (2.96) that of the control group, the intervention clearly improved food intake.

Results also suggest that the intervention had an effect of mitigating weight loss. Mean weight change was negative (-1.5%) among the control group with as much as 36.6% losing more than 2% of baseline body weight, whereas weight change was positive (+0.6%) in the *Gemura* group with only 20.4% losing more than 2% body weight. Considering that the control group had a much shorter average length of stay (4.81 days) than the *Gemura* group (8.35 days), the difference in weight change is dramatic as the loss in the control group occurred in almost half the time. It is conceivable that the control group could have experienced even greater weight loss with longer lengths of stay under similar conditions, with more approaching the generally considered clinically significant 5% body weight loss. Fortunately, weight loss even in the control group was not as severe as other observational findings in Sub-Saharan Africa, namely one multicentre study in South Africa, Kenya, and Ghana with 20.6% of patients having lost more than 5% body weight in hospital,^35^ and another study in Benin in which two-thirds of the sample lost more than 2%.^36^ Comparable hospital weight change findings in Rwanda are not available, so closer context-specific conclusions are more difficult. These findings of relatively small change (most lost less than 2%) in weight over an average length of stay not more than one week in the entire sample are consistent with well documented research showing that body weight is not highly sensitive to large fluctuations in energy intake over the short-term.^37^

Positive nutrition effects of the intervention are further supported by the improved BMI distribution in the *Gemura* group as compared to the control group. Though both groups at baseline had similar BMI distribution, with higher prevalence of malnutrition in the intervention group, the *Gemura* group experienced a decrease in malnourished, an increase in normal weight, and a decrease in combined overweight and obese, indicating convergence toward normal.

Conversely, the control group showed an unfavourable trend of decreasing weight across all categories except combined overweight and obese. Most notably, since study arm was strongly associated with the prevalence of malnutrition, the results indicate that the intervention mitigated hospital malnutrition.

The difference-in-difference analysis however, found little evidence of a significant relationship between the feeding intervention and the weight and BMI changes. The analysis did show that the program had a higher magnitude of effect than other variables on BMI, but overall indicated that there may have been other, unmeasured variables that contributed to the BMI and weight changes. DID analysis may be limited in capturing the nuance of the distribution effects of BMI and weight changes, where convergence toward normal is more desirable than one direction effects.

It was hypothesized that observed improvements in nutrition would lead to better outcomes in patient recovery, as measured by shorter hospital length of stay. Our findings on LOS and patient reported recovery provided limited evidence to support this hypothesis, since LOS was longer in the intervention group than the control group and patient reported recovery was similarly favourable in both groups. Indeed, LOS results from the intervention group were quite unexpected, since secondary routine data on average LOS across hospital wards in the intervention hospitals during the study period indicate LOS much closer to the control hospital results in our study. One reason for the difference could be that our study sample purposively included patients assessed as socioeconomically vulnerable by hospital social workers.

Because LOS had such weak association with study arm, with less observed effect than all other variables we tested including age, disease category, baseline BMI, province, and sex, the LOS differences between groups cannot be primarily attributed to whether patients received the *Gemura* intervention or not. Length of stay is also likely affected by other, unmeasured variables. Though disease category did not appear to contribute significantly to length of stay, disease severity may have played a role, as found in other studies.^38^ Patient characteristics at the intervention sites showed much greater diversity in province of origin than the control group, suggesting that patients could have been transferred for higher levels of care several times before admission and had greater disease severity. In addition, the intervention sites had more patients in diagnosis categories of NCDs, cancers, and “not classified,” potentially requiring longer treatments and stays. In addition, since neither clinical recovery endpoint data nor follow-up data after discharge, the results provide a limited perspective on patient recovery. LOS in particular seems to be more affected by many other factors besides clinical and health outcomes and as such, is an insufficient measure of patient recovery on its own. More investigation is needed to elucidate the effect of the *Gemura* feeding program on patient recovery.

Results on costs of care show that patients spent on average nearly 40% less on food and beverages in the *Gemura* group compared to the control group, with high statistical significance. This difference in average spend (7,305 RWF or about 5.23 USD) represents a considerable reduction in out-of-pocket costs since Rwanda’s GDP per capita is 966 USD, with 52% of the population living beneath the 2.12 USD per day poverty line.^2^ The higher total costs of stay experienced in the control group is consistent with findings on longer length of stay than the intervention group. But, the average cost savings of 1,948 RWF per day in the *Gemura* group is still more than half the daily income for the majority of the population. By reducing out-of-pocket costs, these findings indicate the *Gemura* intervention has the potential to reduce a well-known barrier to healthcare access in Rwanda.

Patient satisfaction survey results showed little difference between groups, were weakly significant, and as a general measure, did not reflect specific perspectives on food provision. Sample sizes, calculated to show significance for the primary outcome, may not have been large enough to show differences between groups.^39^ Our findings provide little information on patient experiences, which is consistent with recent systematic reviews of using NPS in a healthcare setting as a standalone metric.^40^

Though higher on the single metric visual scale (VAS), HRQOL multi-dimensional scores were only marginally better in the control group for mobility, self-care, and usual activities, and slightly better in the *Gemura* group for pain/discomfort, providing little information from the patient perspective. The EuroQoL EQ-5D-3L has been used only a few times in Rwanda, with previous studies also finding statistically significant differences in VAS scores but not in the dimensional measures.^41–42^ More investigation may be needed on patient experience and perceived quality of life as well as the usefulness of this tool in the Rwandan healthcare context. The most notable difference in our study was in the anxiety/depression dimension, with nearly 40% less in the *Gemura* group reporting any problems. This indicates that the intervention has the potential to improve patient perception of their mental health during the hospital stay.

## CONCLUSION

Based on the synthesized results above, the *Gemura* program was effective in improving food intake, mitigating weight loss, reducing hospital malnutrition, improving BMI, and reducing patient-reported anxiety and depression compared to the control group. Low statistical significance of the program effect estimate (DID) highlights the possibility of other, unmeasured factors with a strong effect on BMI and weight within the relatively short length of the average hospital stay. The program was associated with reduced patient costs for food and beverages, but also with longer hospital stays, and slightly higher total out-of-pocket costs of care. Length of stay may not be a sufficient indicator of nutrition-related recovery due to scant association with study arm. Lower or similar HRQOL and patient satisfaction scores along with low statistical significance, may indicate a need for further investigation of patient perceptions of wellbeing.

Overall, improvements in nutrition, mental health, and out-of-pocket patient costs indicate the potential for the *Gemura* hospital feeding program to address nutrition gaps in hospital service delivery, to improve mental health, and to reduce economic barriers to care.

## Data Availability

All data produced in the present study are available upon reasonable request to the authors.

## ACKNOWLEDGEMENTS

The authors would like to thank the staff at Solid’Africa for their contribution and collaboration in carrying out this study, as well as all temporary research assistants and healthcare workers and administrators at each study site. We would also like to thank Solid’Africa’s funding partners who provide unrestricted funding that enables local leaders to own and initiate independent research.

## DATA AVAILABILITY STATEMENT

Data are available upon reasonable request.

## CONTRIBUTORSHIP STATEMENT

SW: project conceptualisation, protocol development, data analysis, data interpretation and preparation of manuscript. EM: protocol development, data analysis, data interpretation and manuscript preparation. MG: data collection, data interpretation and manuscript preparation. LN: project conceptualization, data interpretation and manuscript preparation. SW is the guarantor for this manuscript.

## REFLEXIVITY STATEMENT

**1. How does this study address local research and policy priorities?**

This study is specifically designed to address key questions of local Rwandan organization Solid’Africa as well as their local implementing partners about the effectiveness of their intervention and about the needs of hospital inpatients in Rwanda.

**2. How were local researchers involved in study design?**

The first category of local researchers involved were Solid’Africa program staff (MG and LN) with experience of local data collection and analysis, crucial insight and information about the intervention, relationships with local hospital program implementation sites, and the ability to guide and direct temporary research assistants in data collection. Their contributions were important for study design practical aspects, data collection and cleaning, and interpretation. The second category was a local researcher affiliated with the University of Rwanda (EM) with statistics experience and participation in several studies in Rwanda plus experience with the Rwanda National Ethics Committee. EM contributed to methodology, sampling, analysis, and interpretation. There was one high-income country researcher (SW) with prior research experience in Africa and extensive health program implementation experience in LMICs. Her contribution as a third-party researcher was to provide external study design leadership, coordinating the study and making contributions in research questions, methods, analysis, interpretation, and reporting.

**3. How has funding been used to support the local research team?**

Full-time staff of Solid’Africa were paid salaries and their research duties were included in their job duties. Temporary research assistants were paid a stipend during the time they carried out data collection. External local research team members were paid fair market-based fees for their work.

**4. How are research staff who conducted data collection acknowledged?**

Solid’Africa staff responsible for data collection were included as authors and additional data collectors were acknowledged in the relevant section.

**5. Do all members of the research partnership have access to study data?**

All members of the partnership have access to data.

**6. How was data used to develop analytical skills within the partnership?**

Data review sessions were held within the research team including open discussion about interpretation of findings. Statistical analysis was documented in detail and provided to all research team members.

**7. How have research partners collaborated in interpreting study data?**

Study members have participated in data analysis and interpretation open discussions both virtually and in person. In addition, results were presented in dissemination meetings with research partners from each study site as well as with the Solid’Africa leadership and program teams. Their comments and questions were considered in discussion sections and presentation of results.

**8. How were research partners supported to develop writing skills?**

All research partners were consulted and contributed to draft reviews.

**9. How will research products be shared to address local needs?**

This study will be published as open access. Dissemination of this study and other reports including results will be prioritized as part of Solid’Africa’s communications plans.

**10. How is the leadership, contribution and ownership of this work by LMIC researchers recognised within the authorship?**

SW developed the manuscript incorporating results reports developed by EM who has been recognized with second authorship. Solid’Africa research ownership and leadership contribution was acknowledged with last authorship by LN.

**11. How have early career researchers across the partnership been included within the authorship team?**

We have included early career researchers (SW, MG and LN) within the authorship team.

**12. How has gender balance been addressed within the authorship?**

Three authors are female (SW, MG, and LN) and one author is male (EM).

**13. How has the project contributed to training of LMIC researchers?**

Two Rwandan research team members (MG and LN) are early career researchers and gained first-time experience on aspects of the research process. In addition, many of the research assistants and other data collectors experienced first-time research processes and activities.

**14. How has the project contributed to improvements in local infrastructure?**

This project has not directly contributed to improvements in local infrastructure.

**15. What safeguarding procedures were used to protect local study participants and researchers?**

We have abided by generally accepted and locally approved ethics procedures to safeguard participants’ identities, rights, and privacy.

## REFERENCES

1. United Nations. The 17 Goals [online]. 2023. Available from: https://sdgs.un.org/goals (Accessed 4 Jan 2024).

2. USAID. Rwanda Country Data. [online]. 2023. Available from: https://idea.usaid.gov/cd/rwanda/ (Accessed 21 Dec 2023).

3. Velasco C, Garcia E, Rodriguez V, Frias L, Garriga R, Alvarez J, et al. Comparison of four nutritional screening tools to detect nutritional risk in hospitalized patients: A multicentre study. Eur J Clin Nutr. 2011;65:269–74. doi: 10.1038/ejcn.2010.243

4. Correia MITD, Waitzberg DANL. The impact of malnutrition on morbidity, mortality, length of hospital stay and costs evaluated through a multivariate model analysis. Clin Nutr. 2003;22:235–9. doi: 10.1016/s0261-5614(02)00215-7

5. Sorensen J, Kondrup J, Prokopowicz J, Schiesser M, Krähenbühl L, Meier R, et al. EuroOOPS: An international, multicentre study to implement nutritional risk screening and evaluate clinical outcome. Clin Nutr. 2008;27(3):340–9. doi: 10.1016/j.clnu.2008.03.012

6. Schiesser M, Muller S, Kirchhoff P, Breitenstein S, Schafer M, Clavien P. Assessment of a novel screening score for nutritional risk in predicting complications in gastro-intestinal surgery. Clin Nutr. 2008;27(4):565–70. doi: 10.1016/j.clnu.2008.01.010

7. Alvarez-Hernandez J, Villa M, Leon-Sanz M, De Lorenzo A, Celaya-Perez S. Prevalence and costs of malnutrition in hospitalized patients: The PREDyCES Study. Nutr Hosp. 2012;27:1049–59. doi: 10.3305/nh.2012.27.4.5986

8. Amaral TF, Antunes A, Cabral S, Alves P. An evaluation of three nutritional screening tools in a Portuguese oncology centre. J Hum Nutr Diet. 2008;21:575–83. doi: 10.1111/j.1365-277X.2008.00917.x

9. Ozkalkanli MY, Ozkalkanli DT, Katircioglu K, Savaci S. Comparison of tools for nutrition assessment and screening for predicting the development of complications in orthopedic surgery. Nutr Clin Pract. 2009;24(2):274–80. doi: 10.1177/0884533609332087

10. Lim SL, Ong KCB, Chan YH, Loke WC, Ferguson M, Daniels L. Malnutrition and its impact on cost of hospitalization, length of stay, readmission and 3-year mortality. Clin Nutr. 2012;31(3):345–50. doi: 10.1016/j.clnu.2011.11.001

11. Tangvik RJ, Tell GS, Eisman JA, Berit A, Henriksen A, Miodini R, et al. The nutritional strategy: Four questions predict morbidity, mortality and health care costs. Clin Nutr. 2014;33(4):634–41. doi: 10.1016/j.clnu.2013.09.008

12. Khalatbari-Soltani SMV, P. Impact of nutritional risk screening in hospitalized patients on management, outcome and costs: A retrospective study. Clin Nutr. 2016;35(6):1340–6. doi: 10.1016/j.clnu.2016.02.012

13. Asiimwe SB, Muzoora C, Wilson LA, Moore CC. Bedside measures of malnutrition and association with mortality in hospitalized adults. Clin Nutr. 2015;34(2):252–6. doi: 10.1016/j.clnu.2014.03.013

14. Weatherspoon D, Miller S, Ngabitsinze JC, Weatherspoon LJ, Oehmke JF. Stunting, food security, markets, and food policy in Rwanda. BMC Public Health. 2019;19:882. doi: 10.1186/s12889-019-7208-0

15. USAID. Rwanda: Nutrition Profile 2017-2020. [online]. 2018. Available from: https://2017-2020.usaid.gov/sites/default/files/documents/1864/Rwanda-Nutrition-Profile-Mar2018-508.pdf (Accessed 21 Dec 2023).

16. Wischmeyer PE. Optimising nutrition for recovery after ICU. ICU Management & Practice. 2017;17(3):156–8. Available from: https://healthmanagement.org/c/icu/issuearticle/optimising-nutrition-for-recovery-after-icu

17. Moisey LL, Merriweather JL, Drover JW. The role of nutrition rehabilitation in the recovery of survivors of critical illness: underrecognized and underappreciated. Crit Care. 2022;26:270. doi: 10.1186/s13054-022-04143-5

18. Fearon KCH, Luff R. The nutritional management of surgical patients: enhanced recovery after surgery. Proc Nutr Soc. 2003;62:807–11. doi: 10.1079/PNS2003299

19. Tappenden KA, Quatrara B, Parkhurst ML, Malone AM, Fanjiang G, Ziegler TR. Critical role of nutrition in improving quality of care: An interdisciplinary call to action to address adult hospital malnutrition. J Acad Nutr Diet. 2013;113:1219–37. doi: 10.1016/j.jand.2013.05.015

20. Caesar MP, Ziegler TR. Nutritional support in clinical illness and recovery. Lancet Diabetes Endocrinol. 2015;3(9):734–45. doi: 10.1016/S2213-8587(15)00222-3

21. Nii M, Maeda K, Wakabayashi H, Nishioka S, Tanaka A. Nutritional improvement and energy intake are associated with functional recovery in patients after cerebrovascular disorders. J Stroke Cerebrovasc Dis. 2016;25(1):57–62. doi: 10.1016/j.jstrokecerebrovasdis.2015.08.033

22. Bear DE, Wandrag L, Merriweather JL, Connolly B, Hart N, Grocott MPW, ERACIP Investigators. The role of nutritional support in the physical and functional recovery of critically ill patients: a narrative review. Crit Care. 2017;21:226. doi: 10.1186/s13054-017-1810-2

23. Seneza C, McIsaac DI, Twagirumugabe T, Bould MD. A prospective, cohort study of the effect of acute and chronic malnutrition on length of stay in children having surgery in Rwanda. Anesth Analg. 2022;135(1):152–8. doi: 10.1213/ANE.0000000000005956

24. Nardell M. Nutritional Status of Pediatric Inpatients in Kigali, Rwanda. Yale Medicine Thesis Digital Library, 2014. Available from: http://elischolar.library.yale.edu/ymtdl/1909 (Accessed 10 January 2024).

25. Abahuje E, Niyongombwa, I., Karenzi, D., Bismimwa, J., Tuyishime, E. Ntirenganya, F., Rickard, J. Malnutrition in Acute Care Surgery Patients in Rwanda. World J Surg 2020;44(5):1361–67. doi: 10.1007/s00268-019-05355-7

26. Bianco J. Nutritional Status Among Critically Ill Patients in Rwanda. Mount Saint Vincent University Graduate Theses, 2022. Available from: https://ec.msvu.ca/items/f6dea5cd-08ca-4187-a2d4-34064c9605fc (Accessed 10 January 2024).

27. The East African. Patients decry cost of meals as hospitals adopt feeding policy [online]. 2017. Available from: https://www.theeastafrican.co.ke/tea/rwanda-today/news/patients-decry-cost-of-meals-as-hospitals-adopt-feeding-policy-1364052 (Accessed 4 January 2024).

28. Cederholm T, Bosaeus I, Barazzoni R, Bauer J, Van Gossum A, Klek S, Muscaritoli M, Nyulasi I, Ockenga J, Schneider SM, de van der Schueren MA, Singer P. Diagnostic criteria for malnutrition - An ESPEN Consensus Statement. Clin Nutr. 2015 Jun;34(3):335–40. doi: 10.1016/j.clnu.2015.03.001

29. EuroQol Group. EQ-5D-3L. [online]. 2024. Rotterdam: EuroQol Research Foundation. Available from: https://euroqol.org/information-and-support/euroqol-instruments/eq-5d-3l/ (Accessed 11 January 2024).

30. EuroQol. EQ-5D-5L | About. [online]. 2024. Available from: https://euroqol.org/eq-5d-instruments/eq-5d-5l-about/ (Accessed 11 January 2024).

31. Reichheld FF. The One Number You Need to Grow. [online]. Harvard Business Review. Cambridge: Harvard Business Publishing, 2003:46–54. Available from: https://hbr.org/2003/12/the-one-number-you-need-to-grow (Accessed 17 January 2024).

32. National Health Service England. Using the Friends and Family Test to improve patient experience. [online]. 2020. Available from: https://www.england.nhs.uk/fft/fft-guidance/ (Accessed 10 January 2024).

33. Ogrinc G, Davies L, Goodman D, Batalden P, Davidoff F, Stevens D. SQUIRE 2.0 (Standards for QUality Improvement Reporting Excellence): revised publication guidelines from a detailed consensus process. BMJ Qual Saf. 2016 Dec;25(12):986–992. doi: 10.1136/bmjqs-2015-004411

34. Baquero, A. Net Promoter Score (NPS) and Customer Satisfaction: Relationship and Efficient Management. Sustainability, 2022;14:2011. doi: 10.3390/su14042011

35. Blaaw R, Achar, E., Dolman, R.C., Harbron, J., Moens, M., Munyi, F., Nyatefe, D., Visser, J. The Problemn of Hospital Malnutrition in the African Continent. Nutrients 2019;11(9):2028. doi: 10.3390/nu11092028

36. Alassani AC; Hodonou, A.M.; Donvonou, A.C.; Gbessi, G.D.; Ahoui, S.; Dossou, F.M.; Mêhinto, D.K. Frequency and determinants of post-operative undernutrition in patients undergoing visceral surgery at the University Hospital Centre Koutoucou Hubert Maga, Cotonou. Pan African Medical Journal 2018;29(19). doi: 10.11604/pamj.2018.29.19.10805

37. Chow CC and Hall, KD. Short and long-term energy intake patterns and their implications for human body weight regulation. Physiol Behav. 2014;134:60–65. doi: 10.1016/j.physbeh.2014.02.044

38. Eksandari M, Alizadeh Bahmani, AH, Mardani-Fard, HA et al. Evaluation of factors that influenced the length of hospital stay using data mining techniques. BMC Med Inform Decis Mak. 2022;22:280. doi: 10.1186/s12911-022-02027-w

39. Qualtrics. What is a good Net Promoter Score? [online]. 2025. Available from: https://www.qualtrics.com/experience-management/customer/good-net-promoter-score/ (Accessed 8 February 2025).

40. Adams C, Walpola R, Schembri AM, Harrison R. The ultimate question? Evaluating the use of Net Promoter Score in healthcare: A systematic review. Health Expect. 2022 Oct;25(5):2328–2339. doi: 10.1111/hex.13577

41. Hitimana, R., Lindholm, L., Krantz, G. et al. Health-related quality of life determinants among Rwandan women after delivery: does antenatal care utilization matter? A cross-sectional study. J Health Popul Nutr. 2018;37(12). doi: 10.1186/s41043-018-0142-4

42. Allen Ingabire JC, Tumusiime DK, Sagahutu JB, Urimubenshi G, Bucyibaruta G, Pilusa S and Stewart A. Quality of life of survivors following road traffic orthopaedic injuries in Rwanda. Front. Public Health. 2024;12:1405697. doi: 10.3389/fpubh.2024.1405697

